# Adverse events connected to breast cancer treatment and their associated decrease in quality of life scores

**DOI:** 10.1101/2022.11.28.22282806

**Authors:** Katharina Diernberger, Ewan Gray, Marek Atter, Alistair Bullen, Peter Hall

## Abstract

**Background:** Breast cancer is the most commonly diagnosed form of cancer in the UK, with over 55,000 newly diagnosed cases annually. Fortunately, many patients are cured, with a five-year survival rate of about 80%. Adjuvant chemotherapy in early breast cancer is common and has been shown to increase survival but frequently comes with several adverse events. These can impact patients’ quality of life (QoL) and influence health care costs. Relatively little is known about the magnitude of effects on the QoL of specific toxicities and toxicity profiles.

**Methods:** Adverse event and QoL data (using EQ-VAS and EQ5D) from sub-studies embedded in two different randomized controlled trials (RCTs) of standard adjuvant chemotherapy regimens were used in the analysis. Adverse events were grouped into 20 main toxicity categories. QoL data were reported at baseline and following phases of chemotherapy treatment. Correlations between toxicity groups were explored. Univariate and multivariate analyses investigated the association between individual adverse events and reported QoL. To predict the impact of specific adverse events, a regression model specification was developed based on data from one trial using a backwards selection procedure and assessed for validity using data from the other trial.

**Results:** The most frequently reported toxicities in both trials were Alopecia, Lethargy-Depression-Anxiety, Nausea-Vomiting and Stomatitis. The univariate analysis showed a clear decrease in patients’ QoL measured through the visual analogue scale (EQ-VAS). Results based on EQ5D measurements did not show a clear direction of toxicities’ influence on patients’ QoL. Multivariate results demonstrated a significant change in QoL for Lethargy-Depression-Anxiety, Diarrhoea, Skin-disorders, Infection, Dyspnoea-Respiratory and specified pain.

**Conclusions:** Only a small part of the change in patients’ QoL is induced by the different adverse events the patients faced. Results based on VAS showed a much bigger influence of certain adverse events on patients’ QoL than those derived from EQ5D, leading to the question of whether EQ5D’s domains are capturing what is of importance to patients during chemotherapy treatment.

## Introduction

Breast cancer is the most commonly diagnosed form of cancer, with 55,920 newly diagnosed cases and a mortality figure of 11,499 patients when looking at the UK average between 2016 and 2018. (1) It is a comparatively curable disease, with a five-year survival rate of about 80% and a ten-year survival rate of 76% based on the English breast cancer population. (3) The UK aims for a short waiting time between diagnosis and treatment start and introduced a “two-week wait” standard. (2,3) Of those patients diagnosed, a large majority (81%) have surgery to remove the tumour, 63% undergo radiotherapy and 34% receive chemotherapy as part of the primary treatment. (3) The decision to undergo chemotherapy requires careful consideration of the potential harms relative to incremental gains in survival. In addition to the introduction of a host of treatment-induced toxicities, adjuvant therapies such as chemotherapy have a big impact on healthcare costs and patients’ quality of life (QoL). This potentially applies both to patients undergoing treatment, and cancer survivors. (4)

Reliable estimates of the effects of toxicities on preference-based QoL measures such as the EQ-VAS and EQ5D would be invaluable to cost-effectiveness analyses of current and new therapies in the context of modern frameworks of Health Technology Assessment that rely on measuring costs per Quality Adjusted Life Years (QALYs) gained. A lack of such data has resulted in significant uncertainty in recent value-based decision-making on new cancer therapies. (5–7)

QALYs combine estimates of patient utility with life years to produce a composite measure which captures both elements. Existing research suggests that utilities are affected by the adverse events connected to chemotherapy treatment. (8) Adverse events can be limited to the treatment period or long-lasting, can necessitate dose changes, lead to a discontinuation of treatment and/or may result in patient deaths. (9–12) Nevertheless, too little is known about how specific toxicities and toxicity profiles influence utility and their impact on the QoL of patients during and after treatment. Few clinical trials of adjuvant therapies have directly captured utility values connected to toxicities. The availability of toxicity-specific utility values is crucial in estimating the QALY implications of new adjuvant therapies and potentially helpful in assessing patients’ QoL.

As toxicities and their frequency within specific chemotherapy regimens are usually captured in clinical trials, it theoretically is possible to build algorithms that predict utilities directly from toxicity profiles, provided quality-of-life data has been captured in parallel. Suitable QoL data is not frequently available from historical trials that form our current evidence base for adjuvant chemotherapy.

This study took advantage of the unique clinical trial datasets of the TACT and TACT2 trials that captured relevant QoL data in parallel with high-quality toxicity data. (13,14)

## Aim and research questions

The study’s main aim was to create a user-friendly chemotherapy-toxicity-utility-lexicon of adverse events (AEs) and their associated decrease in QoL scores.

Further, this study has been designed to test the following hypothesis: A higher maximum level of severity of an adverse event is connected to lower QoL outcomes for breast cancer patients receiving adjuvant chemotherapy. There are several connected questions, which we aimed to answer in the course of this analysis.

i. Is the influence of an AE on the QoL dependent on the maximum grade of severity patients experience an adverse event?
ii. Are there correlations between different adverse events?
iii. Is there any variation between the types of adverse events when measuring the impact on patients’ quality of life?
iv. Does the impact of specific AEs on patients’ quality of life change during the study period?

## Methods

### Data

Data from two different randomized controlled trials (RCTs) of standard adjuvant chemotherapy regimens were used. Data from the TACT2 trial was utilised for model development and data from TACT for validating the model.

### Background information on the clinical trials TACT and TACT2

The TACT2 trial enrolled 1238 patients with surgically treated early-stage breast cancer who underwent adjuvant chemotherapy in a quality-of-life sub-study. (14) Early-stage disease was defined by a tumour size of 0–3cm, zero to 10 Nodes involved and zero Metastases (T 0-3, N 0-2, M 0). Additional inclusion criteria determined that patients were 18 years or older and were either fully active and uninhibited or restricted only in physically strenuous activity. (3) The TACT2 study was split into four different treatment arms:

- 4-cycles of epirubicin every 3 weeks followed by 4-cycles of Classical/Bonadonna CMF
- 4-cycles of epirubicin every 3 weeks followed by 4-cycles of oral capecitabine
- 4-cycles of epirubicin every 2 weeks with pegylated GCSF support followed by 4-cycles of Classical/Bonadonna CMF
- 4-cycles of epirubicin every 2 weeks with pegylated GCSF support followed by 4-cycles of oral capecitabine

Participants were asked to complete health status questionnaires at different follow-up time points. The timing of data collection was dependent on the allocated treatment arm and determined by the variation in the cycle length of the prescribed chemotherapy. Assessment time-points were split into “baseline” ie prior to the first chemotherapy administration, “phase 1” ie after the first four cycles and “phase 2” ie after finishing the later four cycles. The trial design allowed for some flexibility around the exact treatment days depending on patients’ clinical presentations, hence some patients had their last follow-up in week 24.

TACT2 results suggest accelerated epirubicin improves outcomes compared to standard epirubicin, and capecitabine gives similar efficacy but a milder side-effect profile to standard CMF. (8) Furthermore, serious Common Terminology Criteria for Adverse Events (CTCAE) graded toxicity was less common with accelerated epirubicin compared to non-accelerated epirubicin. (15) However, patients reported poorer global-QoL, fatigue and role function with CMF compared to oral Capecitabine which does not appear to be explained by the impact on daily activities due to individual side effects. (14)

Ultimately, however, no statistically significant difference in QoL was found between the two groups (accelerated versus non-accelerated), supporting the implementation of a recent meta-analysis that suggested a moderated reduction in 10-year recurrence risk with accelerated regimens. (16)

A similar trial to TACT2 is its predecessor, TACT. Ellis *et al* published results from the study in 2009, exhibiting no overall gain from using docetaxel over standard anthracycline chemotherapy. (17) The full TACT study involved 4123 patients from 103 UK centres and 1 Belgium cancer centre. The QoL sub-study included a sub-sample of 800 patients. Patients were randomised to the treatment group: 4 cycles of FEC followed by 4 cycles of docetaxel, or the control group: the standard treatment at that time, 8 cycles of FEC or 4 cycles of epirubicin followed by 4 cycles of CMF. Results suggested that the addition of a taxane to standard chemotherapy gave no survival benefit (p=0.25) over usual chemotherapy. (13)

### Data preparation

All non-haematological adverse events were grouped into 20 main toxicity categories that aligned with the dominant toxicities reported in the TACT and TACT2 trials. The remaining adverse events were collapsed into an “*other”* variable. The remaining groups were: *Alopecia, Lethargy-Depression-Anxiety, Nausea & Vomiting, Stomatitis, Constipation, Diarrhoea, Neuropathy, Skin disorders, Dyspepsia & Dysgeusia, Myalgia & Arthralgia, Infection, Nail disorders, Pyrexia, Oedema, Eye disorders, Dyspnoea & Respiratory, Pharynx-Larynx-Nasal disorders, Vein disorders* and *Pain-specified*, with the latter including several of the original categories ie *pain in extremity, headache, back pain, musculoskeletal pain, migraine, breast pain, joint stiffness, bone pain, abdominal pain* and *upper abdominal pain*. The full list of diagnosis and their correlating grouping can be found in **Error! Reference source not found**. and **Error! Reference source not found**..

EQ5D utility weights were calculated based on the English value set. EQ-VAS values were included for baseline and at the end-points of phases 1 and 2. Phase 1 includes chemotherapy cycles 1 to 4 and phase 2 includes all remaining cycles and usually ended after the 7^th^ or 8^th^ therapy cycle. For each patient and study phase, AEs are recorded at different levels of severity. Multiple records of an AE within a single phase are possible. We selected the maximum severity each AE was recorded at within each phase as representative of the impact of the AE on QoL. The resulting data structure includes one observation per phase per individual with the maximum severity of each AE experienced in that phase.

### Statistical Analysis

Spearman correlations for the TACT and TACT2 studies between toxicity groups were examined (**Error! Reference source not found**. & **Error! Reference source not found**.). The maximum CTCAE graded toxicity experienced in each period was categorised as “*none”* for grade 0, *Mild/Moderate (MM)* for toxicity levels one and two and *Severe/Life-threatening (SV)* for a toxicity event experienced at level two or above. AEs without grading were excluded from the analysis.

Univariate Analysis based on a linear regression model examined the association between the toxicities and their connected change in utility values. Analysis was repeated on three different samples, using data from the whole trial period, as well as separately for phase one and phase 2. Multivariate analysis included all toxicities in the linear regression model. Two specifications were explored. Firstly, including all levels of adverse events and secondly, including only severe/life-threatening toxicities.

Alternative strategies to account for the influence of missing data were explored. These included complete case analysis, mean imputation and multiple imputation using chained equations. (18)

A final model was developed based on TACT2 data using backwards selection and included all previously identified AEs with a p-value smaller than 0.05. This model was assessed for validity in the TACT data, by which we mean the same toxicities reached statistical significance (p < 0.05).

Data was received in excel and transferred via Datasync hosted by the University of Edinburgh. The analysis was conducted using STATA 14.

## Results

Table 1 presents the frequency of the overall occurrence of grouped toxicities including mild/moderate and severe/life-threatening adverse events. It includes data from all patients enrolled in TACT2 (N=1238) as well as data from all of the primarily enrolled patients from TACT (N=4162 randomised; N=4135 eligible). Some of the exclusion criteria applied within the statistical analysis of the original TACT paper (such as early discontinuation of treatment) were not relevant to this work; therefore numbers are not strictly aligned. This table shows all adverse events for the patient cohort up to patients’ discontinuation. The most frequently reported toxicities in TACT and TACT2 were *Alopecia, Lethargy-Depression-Anxiety, Nausea-Vomiting and Stomatitis*, whereby *Lethargy-Depression-Anxiety* were most commonly experienced in the severe form.

**Table 1:** TACT2 and TACT toxicity frequency divided into mild/moderate and severe with corresponding %-values

| TACT2 AE frequ.<br>(N= 1238) | TACT2 |  |  |  |  |  | TACT |  |  |  |  |  |
| --- | --- | --- | --- | --- | --- | --- | --- | --- | --- | --- | --- | --- |
|  | AE | % | MM | % | SV | % | AE | % | MM | % | SV | % |
| All | **1238** |  |  |  |  |  | **4135** |  |  |  |  |  |
| Alopecia | **1151** | 92.99 | **1151** | 92.99 | **0** | 0.00 | **4036** | 97.61 | **3748** | 90.65 | **288** | 6.97 |
| Lethargy_Depression_Anxiety | **1094** | 88.35 | **1019** | 82.29 | **75** | 6.06 | **3578** | 86.53 | **3029** | 73.26 | **549** | 13.28 |
| Nausea_Vomiting | **852** | 68.79 | **815** | 65.87 | **36** | 2.92 | **2891** | 69.91 | **2647** | 64.01 | **244** | 5.90 |
| Stomatitis | **830** | 67.03 | **790** | 63.83 | **40** | 3.20 | **2201** | 53.23 | **2045** | 49.46 | **156** | 3.77 |
| Constipation | **586** | 47.35 | **579** | 46.76 | **7** | 0.60 | **1790** | 43.30 | **1745** | 42.19 | **46** | 1.11 |
| Diarrhoea | **572** | 46.18 | **519** | 41.93 | **53** | 4.25 | **1115** | 26.96 | **1014** | 24.53 | **100** | 2.43 |
| Neuropathy | **198** | 16.03 | **196** | 15.87 | **2** | 0.16 | **934** | 22.59 | **902** | 21.82 | **32** | 0.77 |
| Skin_disorder | **405** | 32.73 | **394** | 31.80 | **11** | 0.93 | **1055** | 25.51 | **1026** | 24.82 | **28** | 0.68 |
| Dyspepsia_Dysgeusia | **317** | 25.62 | **311** | 25.12 | **6** | 0.50 | **693** | 16.75 | **666** | 16.11 | **26** | 0.64 |
| Myalgia_Arthralgia | **105** | 8.51 | **103** | 8.35 | **2** | 0.16 | **754** | 18.24 | **641** | 15.49 | **114** | 2.75 |
| Infection | **144** | 11.61 | **115** | 9.29 | **29** | 2.32 | **762** | 18.43 | **549** | 13.28 | **213** | 5.16 |
| Nail_disorder | **39** | 3.12 | **39** | 3.12 | **0** | 0.00 | **335** | 8.10 | **332** | 8.03 | **3** | 0.06 |
| Concomitants_Temperature | **69** | 5.55 | **68** | 5.49 | **1** | 0.05 | **280** | 6.78 | **266** | 6.44 | **14** | 0.34 |
| Oedema | **49** | 3.93 | **47** | 3.79 | **2** | 0.15 | **278** | 6.73 | **265** | 6.41 | **13** | 0.32 |
| Concomitants_Eye | **172** | 13.88 | **172** | 13.87 | **0** | 0.01 | **530** | 12.81 | **507** | 12.25 | **23** | 0.55 |
| Dyspnoe_Respiratory | **209** | 16.86 | **187** | 15.14 | **21** | 1.72 | **451** | 10.91 | **411** | 9.95 | **40** | 0.96 |
| Pharynx_Larynx_Nares | **154** | 12.43 | **152** | 12.26 | **2** | 0.17 | **270** | 6.52 | **236** | 5.71 | **33** | 0.81 |
| Vein_disorders | **235** | 18.97 | **227** | 18.36 | **8** | 0.61 | **671** | 16.24 | **608** | 14.70 | **63** | 1.53 |
| Pain_specified | **192** | 15.54 | **180** | 14.56 | **12** | 0.98 | **482** | 11.66 | **420** | 10.16 | **62** | 1.49 |
| Palmar_plantarsyndrom* | **352** | 28.40 | **302** | 24.38 | **50** | 4.02 | **1873\*** | 45.30 | **1745** | 42.19 | **129** | 3.11 |
| othertox | **695** | 56.14 | **403** | 32.55 | **60** | 4.82 | **1482** | 35.84 | **1244** | 30.09 | **238** | 5.75 |

AE – Adverse Event; MM – Mild or moderate adverse event; SV – Severe or life-threatening adverse event: \*Palmar plantar syndrome is a commonly observed adverse event solely in TACT2 – this line stands for Mucosal\_inflammation in the TACT study

### Correlations

The highest correlation within the toxicities included in TACT2 was identified between constipation and stomatitis with a coefficient of 0.2285. Just three more correlations above 0.2 were identified. For detailed information see **Error! Reference source not found**. and **Error! Reference source not found**..

### Univariate Analysis

Results from the univariate Analysis including all severe AEs are presented in Figure 1 showing a clear decrease in patients’ QoL measured through the EQ-VAS over the two treatment phases as presented in the right side. More variability was observed in the second phase of the study. Results based on EQ5D measurements were not showing a clear direction of toxicities’ influence on patients’ QoL. Nevertheless, greater variability was again observed in phase 2.

**Figure 1:**
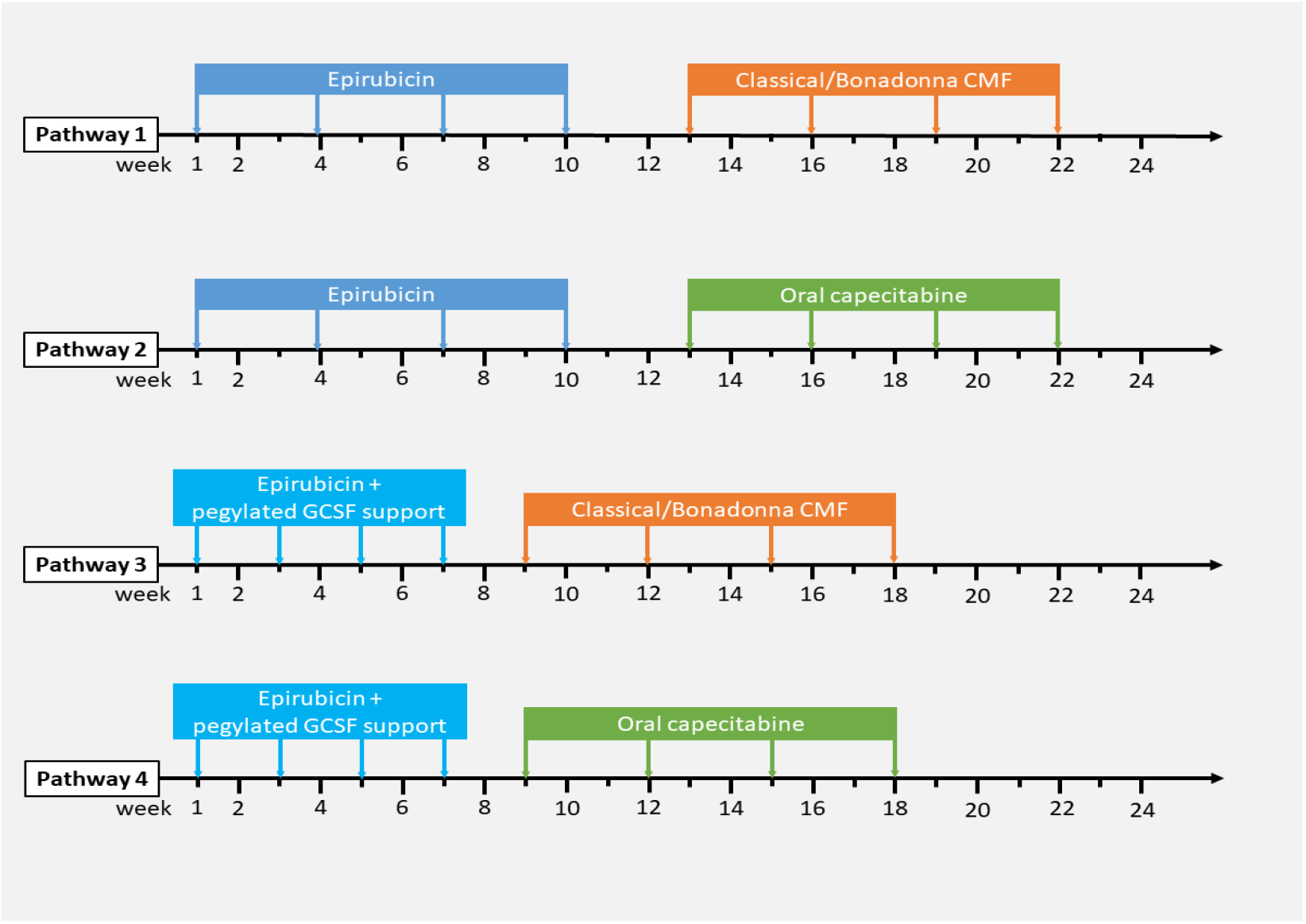
Visual presentation of the four treatment pathways from the TACT2 trial

**Figure 2:**
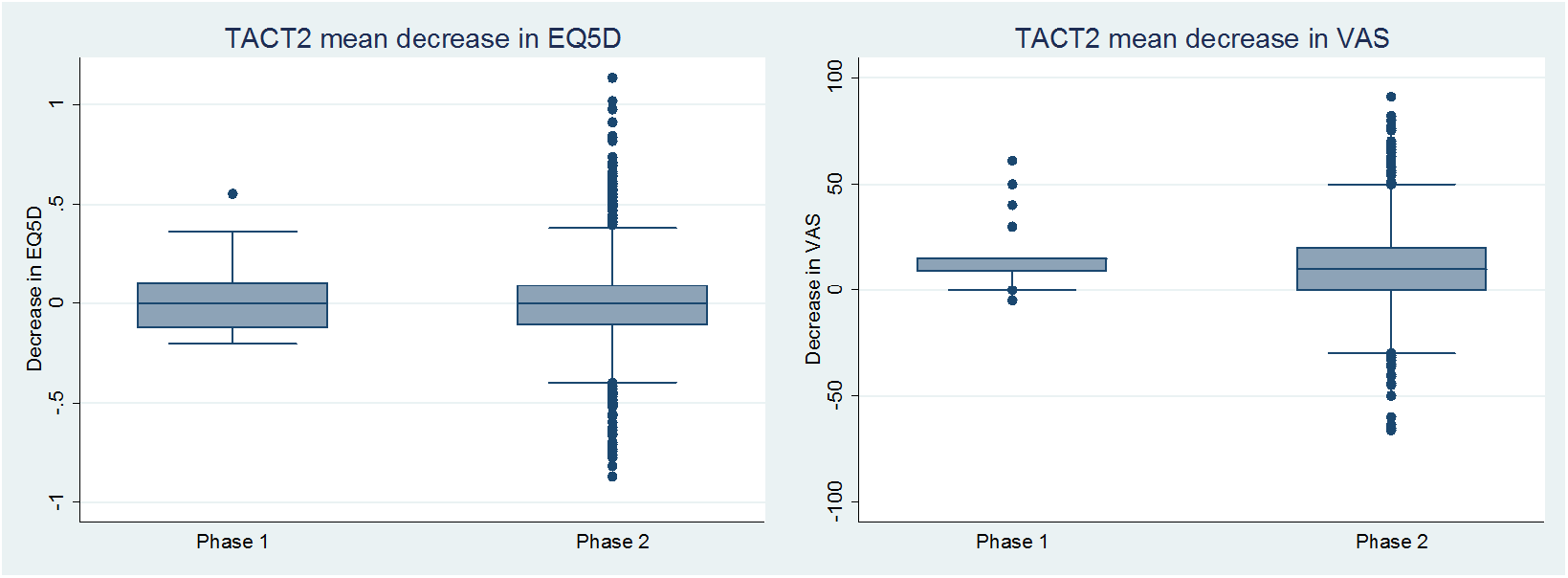
Boxplots of the change in QoL from baseline in EQ5D and EQ-VAS split into “phase1” and “phase2”

### Multivariate Analysis

Results from the multivariate analysis demonstrate the change in QoL. These results are presented for measurements based on EQ5D and EQ-VAS and broken down into phase 1, phase 2 and the overall decrease in QoL. Table 2 presents the multivariate results including toxicities at all levels (including mild/moderate and severe adverse events) and demonstrates a significant change in QoL for the subsequent groups: *Lethargy_Depression_Anxiety, Diarrhoea, Skin_disorders, Infection, Dyspnoe_Respiratory* and *Pain_specified*. Table 3 demonstrates results for severe toxicities only and shows a significant decrease in QoL for some of the toxicities which reached significance in the above analysis (*Lethargy_Depression_Anxiety, Diarrhoea, Skin_disorders* and *Infection*) and for *Oedema and Concomitants_Eye*.

**Table 2:**
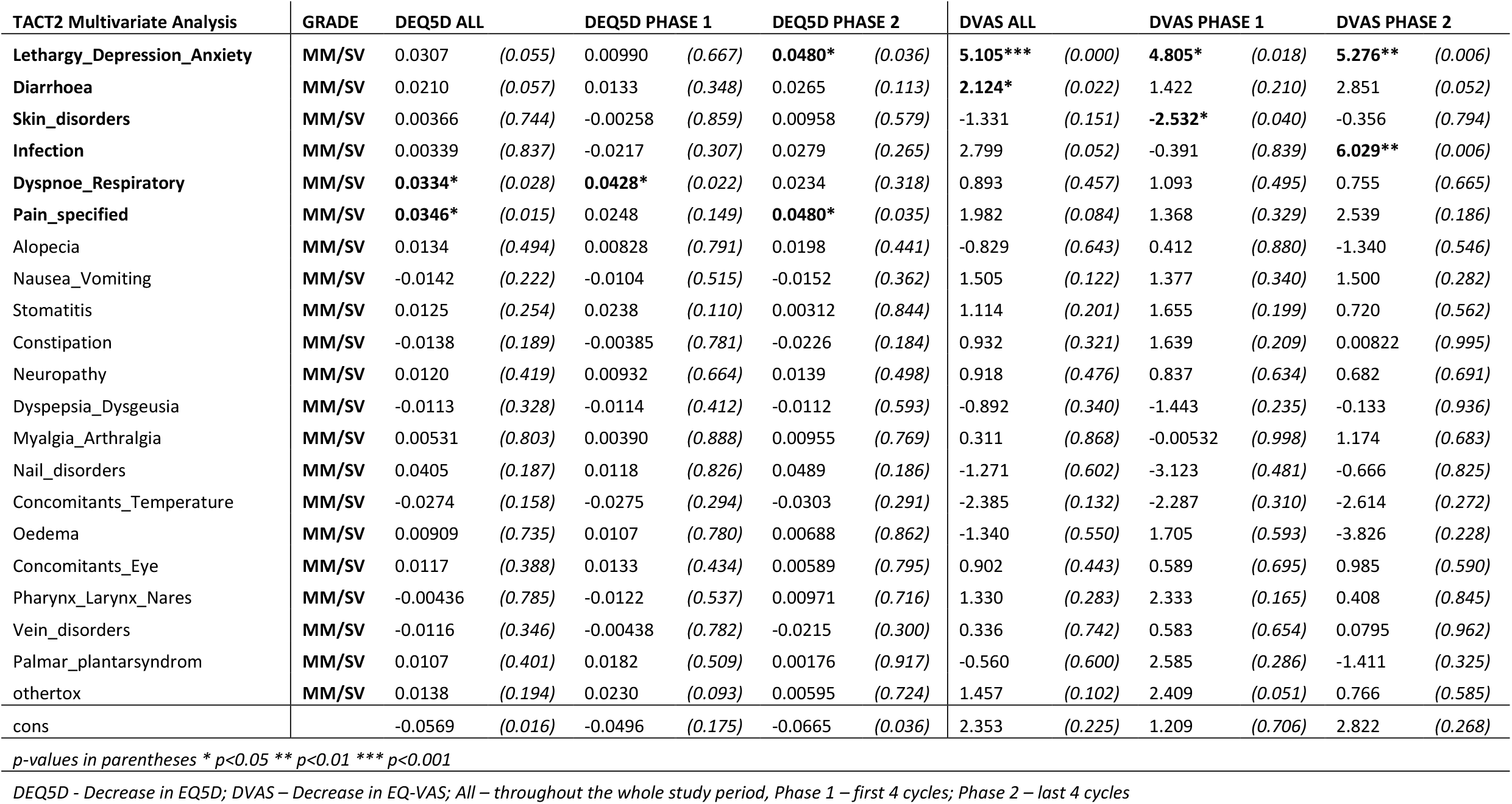
Decrease in quality of life connected to different toxicity profiles experienced at all toxicity levels (MM and SV); results split into decrease measured by EQ5D and EQ-VAS; results presented for the overall study period and split into phase 1 and 2

| TACT2 Multivariate Analysis | GRADE | DEQ5D ALL |  | DEQ5D PHASE 1 |  | DEQ5D PHASE 2 |  | DVAS ALL |  | DVAS PHASE 1 |  | DVAS PHASE 2 |  |
| --- | --- | --- | --- | --- | --- | --- | --- | --- | --- | --- | --- | --- | --- |
| Lethargy_Depression_Anxiety | MM/SV | 0.0307 | (0.055) | 0.00990 | (0.667) | **0.0480\*** | (0.036) | **5.105\*\*\*** | (0.000) | **4.805\*** | (0.018) | **5.276\*\*** | (0.006) |
| Diarrhoea | MM/SV | 0.0210 | (0.057) | 0.0133 | (0.348) | 0.0265 | (0.113) | **2.124\*** | (0.022) | 1.422 | (0.210) | 2.851 | (0.052) |
| Skin_disorders | MM/SV | 0.00366 | (0.744) | -0.00258 | (0.859) | 0.00958 | (0.579) | -1.331 | (0.151) | **-2.532\*** | (0.040) | -0.356 | (0.794) |
| Infection | MM/SV | 0.00339 | (0.837) | -0.0217 | (0.307) | 0.0279 | (0.265) | 2.799 | (0.052) | -0.391 | (0.839) | **6.029\*\*** | (0.006) |
| Dyspnoe_Respiratory | MM/SV | **0.0334\*** | (0.028) | **0.0428\*** | (0.022) | 0.0234 | (0.318) | 0.893 | (0.457) | 1.093 | (0.495) | 0.755 | (0.665) |
| Pain_specified | MM/SV | **0.0346\*** | (0.015) | 0.0248 | (0.149) | **0.0480\*** | (0.035) | 1.982 | (0.084) | 1.368 | (0.329) | 2.539 | (0.186) |
| Alopecia | MM/SV | 0.0134 | (0.494) | 0.00828 | (0.791) | 0.0198 | (0.441) | -0.829 | (0.643) | 0.412 | (0.880) | -1.340 | (0.546) |
| Nausea_Vomiting | MM/SV | -0.0142 | (0.222) | -0.0104 | (0.515) | -0.0152 | (0.362) | 1.505 | (0.122) | 1.377 | (0.340) | 1.500 | (0.282) |
| Stomatitis | MM/SV | 0.0125 | (0.254) | 0.0238 | (0.110) | 0.00312 | (0.844) | 1.114 | (0.201) | 1.655 | (0.199) | 0.720 | (0.562) |
| Constipation | MM/SV | -0.0138 | (0.189) | -0.00385 | (0.781) | -0.0226 | (0.184) | 0.932 | (0.321) | 1.639 | (0.209) | 0.00822 | (0.995) |
| Neuropathy | MM/SV | 0.0120 | (0.419) | 0.00932 | (0.664) | 0.0139 | (0.498) | 0.918 | (0.476) | 0.837 | (0.634) | 0.682 | (0.691) |
| Dyspepsia_Dysgeusia | MM/SV | -0.0113 | (0.328) | -0.0114 | (0.412) | -0.0112 | (0.593) | -0.892 | (0.340) | -1.443 | (0.235) | -0.133 | (0.936) |
| Myalgia_Arthralgia | MM/SV | 0.00531 | (0.803) | 0.00390 | (0.888) | 0.00955 | (0.769) | 0.311 | (0.868) | -0.00532 | (0.998) | 1.174 | (0.683) |
| Nail_disorders | MM/SV | 0.0405 | (0.187) | 0.0118 | (0.826) | 0.0489 | (0.186) | -1.271 | (0.602) | -3.123 | (0.481) | -0.666 | (0.825) |
| Concomitants_Temperature | MM/SV | -0.0274 | (0.158) | -0.0275 | (0.294) | -0.0303 | (0.291) | -2.385 | (0.132) | -2.287 | (0.310) | -2.614 | (0.272) |
| Oedema | MM/SV | 0.00909 | (0.735) | 0.0107 | (0.780) | 0.00688 | (0.862) | -1.340 | (0.550) | 1.705 | (0.593) | -3.826 | (0.228) |
| Concomitants_Eye | MM/SV | 0.0117 | (0.388) | 0.0133 | (0.434) | 0.00589 | (0.795) | 0.902 | (0.443) | 0.589 | (0.695) | 0.985 | (0.590) |
| Pharynx_Larynx_Nares | MM/SV | -0.00436 | (0.785) | -0.0122 | (0.537) | 0.00971 | (0.716) | 1.330 | (0.283) | 2.333 | (0.165) | 0.408 | (0.845) |
| Vein_disorders | MM/SV | -0.0116 | (0.346) | -0.00438 | (0.782) | -0.0215 | (0.300) | 0.336 | (0.742) | 0.583 | (0.654) | 0.0795 | (0.962) |
| Palmar_plantarsyndrom | MM/SV | 0.0107 | (0.401) | 0.0182 | (0.509) | 0.00176 | (0.917) | -0.560 | (0.600) | 2.585 | (0.286) | -1.411 | (0.325) |
| othertox | MM/SV | 0.0138 | (0.194) | 0.0230 | (0.093) | 0.00595 | (0.724) | 1.457 | (0.102) | 2.409 | (0.051) | 0.766 | (0.585) |
| cons |  | -0.0569 | (0.016) | -0.0496 | (0.175) | -0.0665 | (0.036) | 2.353 | (0.225) | 1.209 | (0.706) | 2.822 | (0.268) |

*p-values in parentheses \* p<0.05 \*\* p<0.01 \*\*\* p<0.001*
*DEQ5D - Decrease in EQ5D; DVAS – Decrease in EQ-VAS; All – throughout the whole study period, Phase 1 – first 4 cycles; Phase 2 – last 4 cycles*

**Table 3:** Decrease in quality of life connected to different toxicity profiles on a severe/life-threatening level; results split into decrease measured by EQ5D and EQ-VAS; results presented for the overall study period and split into phases 1 and 2

| TACT2 Multivariate Analysis | GRADE | DEQ5D ALL |  | DEQ5D PHASE 1 |  | DEQ5D PHASE 2 |  | DVAS ALL |  | DVAS PHASE 1 |  | DVAS PHASE 2 |  |
| --- | --- | --- | --- | --- | --- | --- | --- | --- | --- | --- | --- | --- | --- |
| Lethargy_Depression_Anxiety | SV | 0.0439 | (0.086) | 0.000728 | (0.985) | **0.0773\*** | (0.027) | **9.769\*\*\*** | (0.000) | **7.168\*** | (0.036) | **11.36\*\*\*** | (0.000) |
| Diarrhoea | SV | **0.0667\*** | (0.038) | -0.0315 | (0.675) | 0.0753 | (0.058) | 4.837 | (0.117) | -4.456 | (0.476) | 6.425 | (0.113) |
| Skin_disorders | SV | 0.118 | (0.070) | 0.0897 | (0.212) | 0.202 | (0.138) | 5.650 | (0.364) | -2.162 | (0.766) | **26.19\*** | (0.018) |
| Infection | SV | 0.0564 | (0.113) | 0.00233 | (0.963) | **0.102\*** | (0.047) | 4.601 | (0.140) | -0.806 | (0.848) | **9.487\*** | (0.031) |
| Dyspnoe_Respiratory | SV | **0.613\*\*** | (0.006) | 0.690** | (0.001) | 0 | . | 22.59 | (0.349) | 24.70 | (0.312) | 0 | . |
| Pain_specified | SV | -0.0611 | (0.785) | 0 | . | -0.0801 | (0.742) | **-50.51\*\*** | (0.006) | 0 | . | **-45.48\*** | (0.021) |
| Alopecia | SV | 0.0160 | (0.725) | 0.0197 | (0.825) | 0.0115 | (0.835) | -0.533 | (0.888) | 7.797 | (0.341) | -2.897 | (0.514) |
| Nausea_Vomiting | SV | -0.00201 | (0.972) | -0.0158 | (0.831) | 0.00772 | (0.933) | 2.256 | (0.623) | 4.323 | (0.496) | 1.123 | (0.871) |
| Stomatitis | SV | 0.0210 | (0.526) | 0.0396 | (0.372) | 0.0224 | (0.678) | 4.193 | (0.136) | 4.514 | (0.228) | 4.084 | (0.387) |
| Constipation | SV | 0.0315 | (0.407) | 0.00226 | (0.969) | 0.0339 | (0.505) | 3.166 | (0.310) | 7.958 | (0.113) | 0.695 | (0.867) |
| Neuropathy | SV | 0.0167 | (0.850) | 0.0941 | (0.353) | -0.0207 | (0.893) | 2.089 | (0.754) | 9.243 | (0.285) | -2.753 | (0.816) |
| Dyspepsia_Dysgeusia | SV | -0.0591 | (0.665) | -0.0117 | (0.937) | -0.177 | (0.536) | -4.181 | (0.725) | -1.526 | (0.902) | -8.079 | (0.752) |
| Myalgia_Arthralgia | SV | 0.0152 | (0.845) | -0.0721 | (0.468) | 0.105 | (0.401) | -4.816 | (0.510) | -6.384 | (0.444) | -1.301 | (0.920) |
| Nail_disorders | SV | 0.0517 | (0.715) | 0.0878 | (0.520) | 0 | . | 1.970 | (0.865) | 4.720 | (0.680) | 0 | . |
| Concomitants_Temperature | SV | -0.0211 | (0.847) | -0.00209 | (0.989) | -0.0526 | (0.750) | -5.355 | (0.554) | -11.04 | (0.373) | 1.497 | (0.911) |
| Oedema | SV | 0.158 | (0.215) | 0.0505 | (0.811) | 0.252 | (0.129) | 7.042 | (0.539) | 2.588 | (0.910) | 9.970 | (0.458) |
| Concomitants_Eye | SV | 0.0128 | (0.814) | -0.0673 | (0.566) | 0.00906 | (0.891) | 2.766 | (0.534) | 8.378 | (0.385) | -0.165 | (0.976) |
| Pharynx_Larynx_Nares | SV | 0.0254 | (0.332) | -0.0210 | (0.620) | 0.0558 | (0.123) | 3.604 | (0.132) | 4.736 | (0.195) | 4.242 | (0.186) |
| Vein_disorders |  | -0.0578* | (0.015) | -0.0476 | (0.194) | -0.0724* | (0.024) | 2.164 | (0.263) | 1.586 | (0.623) | 2.312 | (0.356) |
| Palmar_plantarsyndrom | SV | 0.0439 | (0.086) | 0.000728 | (0.985) | 0.0773* | (0.027) | 9.769*** | (0.000) | 7.168* | (0.036) | 11.36*** | (0.000) |
| othertox | SV | 0.0667* | (0.038) | -0.0315 | (0.675) | 0.0753 | (0.058) | 4.837 | (0.117) | -4.456 | (0.476) | 6.425 | (0.113) |
| cons |  | 0.118 | (0.070) | 0.0897 | (0.212) | 0.202 | (0.138) | 5.650 | (0.364) | -2.162 | (0.766) | 26.19 | (0.018) |

p-values in parentheses \* p<0.05 \*\* p<0.01 \*\*\* p<0.001
DEQ5D - Decrease in EQ5D; DVAS – Decrease in EQ-VAS; All – throughout the whole study period, Phase 1 – first 4 cycles; Phase 2 – last 4 cycles

In the next step, the eight identified toxicity groups were included in a final multivariate model (backward selection). All of them reached significant values, demonstrating similar decreases in EQ5D and EQ-VAS scores to the wider multivariate models, as can be seen in Table 4.

**Table 4:**
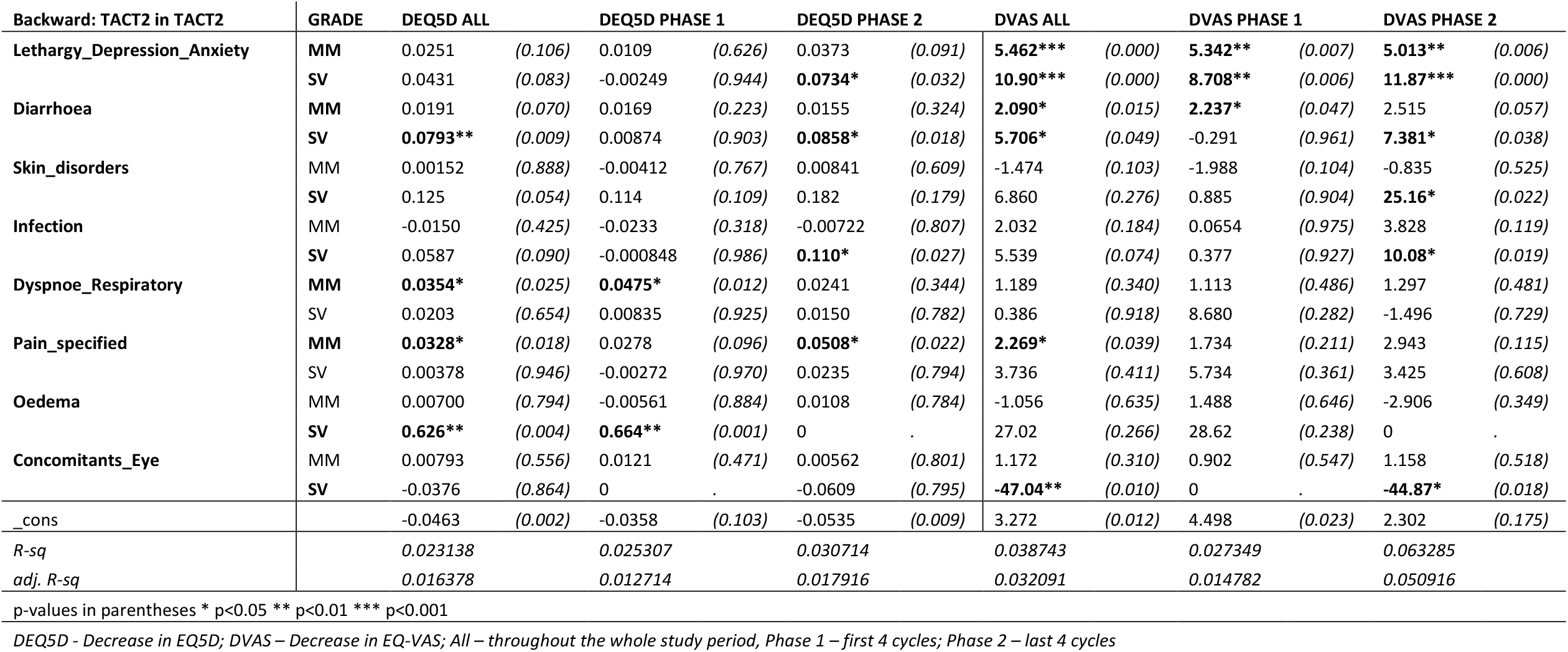
Backward selected TACT2 toxicities – final multivariate model

### Validating the model (generated from TACT2 data) using TACT data

In the validation stage, the final model was fitted to the TACT data (Table 5) demonstrating that out of the eight significant toxicities in TACT2 five demonstrated a significant change in QoL in TACT (bold) with diarrhoea, pain, and eye disorders no longer providing independent prediction.

**Table 5:**
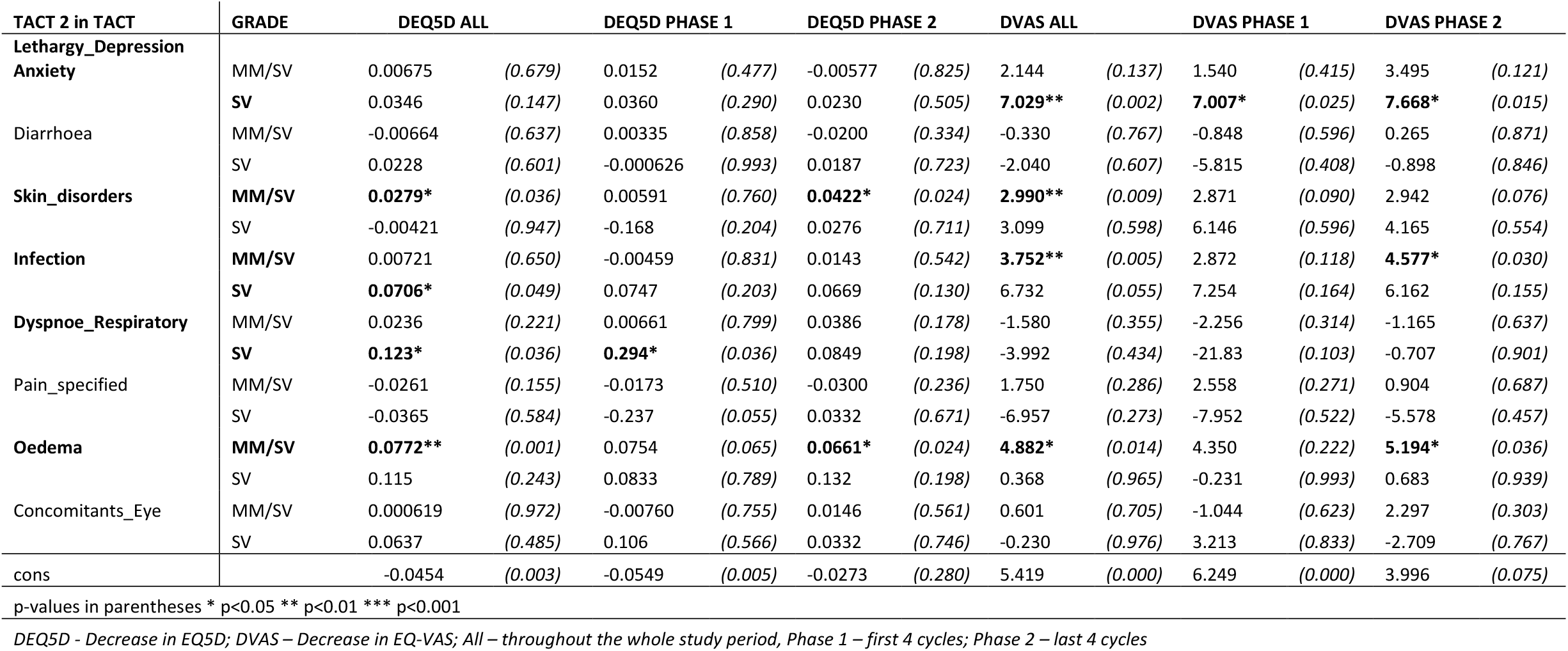
Validation of the multivariate model derived from TACT2, in the TACT dataset; models were run separately including all toxicity grades (MM/SV) and for severe/life-threatening toxicities.

## Discussion

Very little research has been done into estimating the effects of AEs on quality of life measures within cost-effectiveness analysis for early breast cancer. Campbell *et al* (3) highlight the “decrement” of health-related QoL-associated toxicities and explain how the over-reliance on clinical opinion and simple assumptions in sourcing HRQoL estimates for adjuvant therapy economic models highlights the scarcity of relevant data. Consequently, this area of health research requires greater knowledge and understanding, as project outputs potentially are of great value.

Despite being in the setting of metastatic breast cancer Cortes *et al* (19) study of eribulin versus capecitabine for EQ5D is a near match in terms of what is currently available. They conclude that utility values for various metastatic breast cancer stages and chemotherapy-related toxicities are an important component of economic analysis, but are scarcely found in clinical trial data. (19) The authors look at toxicity profiles for AEs for eribulin and capecitabine, and AEs grading in a wider context, which is of interest for comparing and validating findings from TACT2 toxicities. The study asserts metastatic breast cancer is associated with the largest utility decrement and negative effect on QoL compared to other and earlier breast cancer stages. (19)

Campbell *et al’s* ‘Cost-effectiveness of adjuvant chemotherapy for early breast cancer’ is particularly relevant, collating data via a meta-analysis of three large-scale RCTs: the Adjuvant Breast Cancer trial (ABC), the National Epirubicin Adjuvant Trial (NEAT) and Taxotere as Adjuvant Chemotherapy Trial (TACT). Campbell *et al* provides a cost-effectiveness comparison of four types of treatment options (no chemotherapy; first-generation chemotherapy (CMF); second-generation chemotherapy (FEC60 or E-CMF); and third-generation chemotherapy (FEC-D and docetaxel) whereby lifetime costs and QALYs for women with early breast cancer are estimated and compared. (4,20,21) Using a parametric, regression-based survival model, Campbell *et al* conclude different treatment strategies have the potential to be cost-effective depending on the type of patient and their prognoses. (4) New regimes are usually more effective but also more toxic and therefore lead to a higher rate of Adverse Events which subsequently has a negative impact on the health-related QoL. (4)

### Answers to RQs

#### RQ1 & RQ3

In this analysis, as indicated by the R^2^ and adjusted R^2^ in the multivariate model, change in QoL only partially explains the AEs experienced by patients, even if only statistically significant toxicities are included in the model. Thus, toxicities, as measured by the CTCAE system, do not seem to be the dominant predictor of life quality, despite several AEs being associated with a decrease in EQ5D scores. This may before three reasons [1] factors other than AEs are driving QoL during chemotherapy, [2] the EQ5D is insufficiently sensitive to QoL changes associated with AEs or [3] the CTCAE system is not adequately characterizing AEs that affect QoL. We conclude that there is currently insufficient evidence at this stage to use toxicities to predict QoL changes.

#### RQ2

Another interesting observation is that the data did not identify any strong correlation between adverse events; the highest correlations were around 0.2 which does not justify an exclusion. (22) This is surprising given that toxicity rates are thought to be mainly due to the pharmacodynamic and pharmacokinetic factors that are likely to be consistent between AEs within individual patients.

#### RQ4

Interestingly, many of the EQ-VAS-measured toxicity-related QoL decreases show a sizeable difference between the two phases, often with a greater effect during the first phase, measured after the first four cycles. This could be due to several reasons ranging from the fact that patients know what they will be facing and are therefore less frightened, due to previous experience they can react more promptly to AEs or may be due to dose modification or discontinuation in patients experiencing early severe side effects. Moreover, the QoL measures (EQ-VAS and EQ5D) are not necessarily in agreement, as one may not capture the same effects as the other. It is also worth considering that in “Phase2” other factors may influence QoL proportionally more than AEs: if the patient is aware of the therapy response on the tumour, the influence of any physical ailment might be overshadowed. Furthermore, the effect on “Phase2” QoL of an adverse event that already occurred during “Phase1” might be reduced because effective symptom control may already be in place. It further is possible that EQ5D domains are unable to capture relevant factors in this phase.

## Data Availability

Data are not available for sharing.

## Authors’ contributions

KD and PH led the conception and design of the study. KD led the data acquisition and conducted data management and analysis supported by EG. KD anf EG led the data interpretation supported by PH. KD drafted the article which was revised by EG, AB, MA, and PH. All authors critically reviewed and edited the paper and approved the final version to be published.

## Ethics

TACT2 is registered with the ISRCTN, number 68068041, and with ClinicalTrials.gov, number NCT00301925. The study was approved by the Scotland Multi-Research Ethics Committee (MREC 04/MRE00/88) and local research and development offices.

TACT (CRUK01/001) was approved by the national South East Multi-Research Ethics Committee (MREC 00/1/59) and the local ethics committees of all participating centres.

## Acknowledgements and Funding

We thank all members of the TACT and TACT2 study team. We further thank James Golding for comments on an earlier version of the paper.

We thank the patients, their relatives and treating clinicians, and the hospitals who agreed to participate in the TACT2 study. We thank Cancer Research UK for funding the study, Amgen, Pfizer, and Roche, for unrestricted educational grants, and the National Institute for Health Research cancer research networks in England and their equivalent NHS R&D-funded networks in Scotland, Wales, and Northern Ireland for “in-kind” support. Finally, we thank the members of the independent data monitoring committee and the Clinical Trials and Statistics Unit at the Institute of Cancer Research overarching trials steering committee, and the two co-sponsoring organisations, The Institute of Cancer Research and NHS Lothian. Roche supplied free capecitabine and Amgen provided subsidised pegfilgrastim for trial use. The pharmaceutical company partners had the opportunity to review the manuscript to ensure any proprietary information was correct.

The research costs of the TACT trial were met by Cancer Research UK (CRUK 01/001) and by educational grants from Sanofi-Aventis, Pfizer, and Roche. Trial recruitment was facilitated within centres by the NIHR-funded National Cancer Research Network. We thank all the patients who participated in this study, the doctors, nurses, radiographers, physicists, pathologists, and data managers at the participating centres, and the laboratory staff at Royal Marsden Hospital (London, UK), Nottingham City Hospital (Nottingham, UK), and Glasgow Royal Infirmary (Glasgow, UK). Recognition goes to all the trials unit staff at the Information & Statistics Division (ISD), Edinburgh (formerly SCTN), Cancer Trials & Research Unit (CTRU), Leeds, Wales Cancer Trials Network (WCTN), Cardiff, CRUK & UCL Cancer Trials Centre, London and The Institute of Cancer Research’s Clinical Trials & Statistics Unit (ICR-CTSU), Sutton, who contributed to trial management and data collection, with special thanks to M McLinden (ISD), J Copeland (CTRU Leeds), and A Yongue (ICR-CTSU). We would also like to thank the Independent Data Monitoring and Trial Steering Committees for their oversight of the trial and D Mills (ICR-CTSU) for helping to prepare the manuscript.

## Bibliography

1. Cancer Research UK. Breast cancer statistics [Internet]. [cited 2022 Nov 7]. Available from: https://www.cancerresearchuk.org/health-professional/cancer-statistics/statistics-by-cancer-type/breast-cancer

2. Department of Health and Social Care. NHS Constitution for England [Internet]. [cited 2022 Nov 17]. 2021. Available from: https://www.gov.uk/government/publications/the-nhs-constitution-for-england

3. Breast Cancer Now. What is breast cancer? [Internet]. 2022 [cited 2022 Nov 7]. Available from: https://breastcancernow.org/information-support/have-i-got-breast-cancer/what-breast-cancer?gclid=CLa6qam8q8kCFcaVGwodRgICpQ

4. Campbell HE, Epstein D, Bloomfield D, Griffin S, Manca A, Yarnold J, et al. The cost-effectiveness of adjuvant chemotherapy for early breast cancer: A comparison of no chemotherapy and first, second, and third generation regimens for patients with differing prognoses. Eur J Cancer [Internet]. 2011 Nov 1 [cited 2022 Nov 7];47(17):2517–30. Available from: http://www.ejcancer.com/article/S0959804911004230/fulltext

5. National Institute for Health and Care Excellence. Pertuzumab for adjuvant treatment of early HER2-positive breast cancer. 2018 [cited 2022 Nov 7]; Available from: https://www.nice.org.uk/guidance/gid-ta10184/documents/appraisal-consultation-document

6. National Institute for Health and Clinical Excellence. Bevacizumab in combination with a taxane for the first-line treatment of metastatic breast cancer [Internet]. 2010 [cited 2022 Nov 7]. Available from: https://www.nice.org.uk/guidance/ta214/documents/breast-cancer-bevacizumab-in-combination-with-a-taxane-final-appraisal-determination3

7. National Institute for Health and Care Excellence. Early and locally advanced breast cancer: diagnosis and treatment, Clinical guideline [CG80]. 2009 [cited 2017 Nov 7]; Available from: https://www.nice.org.uk/guidance/cg80

8. Ward S, Simpson E, Davis S, Hind D, Rees A, Wilkinson A. Taxanes for the adjuvant treatment of early breast cancer: systematic review and economic evaluation. Health Technol Assess [Internet]. 2007 [cited 2022 Nov 7];11(40). Available from: https://pubmed.ncbi.nlm.nih.gov/17903394/

9. Litton JK, Rugo HS, Ettl J, Hurvitz SA, Gonçalves A, Lee KH, et al. Talazoparib in Patients with Advanced Breast Cancer and a Germline BRCA Mutation. N Engl J Med [Internet]. 2018 Aug 23 [cited 2022 Nov 7];379(8):753–63. Available from: https://pubmed.ncbi.nlm.nih.gov/30110579/

10. Schmid P, Rugo HS, Adams S, Schneeweiss A, Barrios CH, Iwata H, et al. Atezolizumab plus nabpaclitaxel as first-line treatment for unresectable, locally advanced or metastatic triple-negative breast cancer (IMpassion130): updated efficacy results from a randomised, double-blind, placebocontrolled, phase 3 trial. Lancet Oncol [Internet]. 2020 Jan 1 [cited 2022 Nov 7];21(1):44–59. Available from: https://pubmed.ncbi.nlm.nih.gov/31786121/

11. Bardia A, Hurvitz SA, Tolaney SM, Loirat D, Punie K, Oliveira M, et al. Sacituzumab Govitecan in Metastatic Triple-Negative Breast Cancer. N Engl J Med [Internet]. 2021 Apr 22 [cited 2022 Nov 7];384(16):1529–41. Available from: https://pubmed.ncbi.nlm.nih.gov/33882206/

12. Chan J, Adderley H, Alameddine M, Armstrong A, Arundell D, Fox R, et al. Permanent hair loss associated with taxane chemotherapy use in breast cancer: A retrospective survey at two tertiary UK cancer centres. Eur J Cancer Care (Engl) [Internet]. 2020 Dec 22 [cited 2022 Nov 7];30(3):e13395–e13395. Available from: https://europepmc.org/article/MED/33350015

13. Hall E, Cameron D, Waters R, Barrett-Lee P, Ellis P, Russell S, et al. Comparison of patient reported quality of life and impact of treatment side effects experienced with a taxane-containing regimen and standard anthracycline based chemotherapy for early breast cancer: 6year results from the UK TACT trial (CRUK/01/001). Eur J Cancer. 2014 Sep;50(14):2375–89.

14. Cameron D, Morden JP, Canney P, Velikova G, Coleman R, Bartlett J, et al. Accelerated versus standard epirubicin followed by cyclophosphamide, methotrexate, and fluorouracil or capecitabine as adjuvant therapy for breast cancer in the randomised UK TACT2 trial (CRUK/05/19): a multicentre, phase 3, open-label, randomised, controlled trial. Lancet Oncol [Internet]. 2017 Jul 1 [cited 2022 Nov 7];18(7):929–45. Available from: https://pubmed.ncbi.nlm.nih.gov/28600210/

15. Common Terminology Criteria for Adverse Events (CTCAE) [Internet]. [cited 2022 Nov 7]. Available from: https://ctep.cancer.gov/protocoldevelopment/electronic_applications/ctc.htm

16. Gray R, Bradley R, Braybrooke J, Liu Z, Peto R, Davies L, et al. Increasing the dose intensity of chemotherapy by more frequent administration or sequential scheduling: a patient-level meta-analysis of 37 298 women with early breast cancer in 26 randomised trials. Lancet [Internet]. 2019 Apr 6 [cited 2022 Nov 7];393(10179):1440–52. Available from: https://pubmed.ncbi.nlm.nih.gov/30739743/

17. Ellis P, Barrett-Lee P, Johnson L, Cameron D, Wardley A, O’Reilly S, et al. Sequential docetaxel as adjuvant chemotherapy for early breast cancer (TACT): an open-label, phase III, randomised controlled trial. Lancet [Internet]. 2009 [cited 2022 Nov 7];373(9676):1681–92. Available from: https://pubmed.ncbi.nlm.nih.gov/19447249/

18. Rosenbaum PR, Rubin DB. Reducing bias in observational studies using subclassification on the propensity score. J Am Stat Assoc. 1984;79(387):516–24.

19. Cortes J, Hudgens S, Twelves C, Perez EA, Awada A, Yelle L, et al. Health-related quality of life in patients with locally advanced or metastatic breast cancer treated with eribulin mesylate or capecitabine in an open-label randomized phase 3 trial. Breast Cancer Res Treat [Internet]. 2015 Dec 1 [cited 2022 Nov 7];154(3):509–20. Available from: https://mayoclinic.pure.elsevier.com/en/publications/health-related-quality-of-life-in-patients-with-locally-advanced-

20. ISRCTN registry. ISRCTN - ISRCTN68068041: Trial of accelerated adjuvant chemotherapy with capecitabine in early breast cancer [Internet]. 2022 [cited 2022 Nov 7]. Available from: https://www.isrctn.com/ISRCTN68068041

21. Robinson A, Gyrd-Hansen D, Bacon P, Baker R, Pennington M, Donaldson C. Estimating a WTP-based value of a QALY: the “chained” approach. Soc Sci Med [Internet]. 2013 Sep [cited 2022 Nov 7];92:92–104. Available from: https://pubmed.ncbi.nlm.nih.gov/23849283/

22. Taylor R. Interpretation of the Correlation Coefficient: A Basic Review. Journal of Diagnostic Medical Sonography. 1990;6(1):35–9.

