## Supplementary material for "Adverse events connected to breast cancer treatment and their associated decrease in quality of life scores"

### *Supplementary Table 1: Toxicity groups in TACT*

| TACT - toxicity grouping | N= | % | |
|---|---|---|---|
| ALL patients | 4135 | *100* | |
| Alopecia | 4036 | *97.61* | Alopecia |
| Lethargy_Depression_Anxiety | 3578 | *86.53* | Lethargy, Insomnia, Anxiety, Depression, Depressed mood, Dizziness, Vision blurred |
| Nausea_Vomiting | 2891 | *69.91* | Vomiting |
| Stomatitis | 2201 | *53.23* | Stomatitis, Dry mouth, Oral candidiasis, Oral herpes |
| Constipation | 1790 | *43.30* | Constipation, Haemorrhoids |
| Diarrhoea | 1115 | *26.96* | Diarrhoea |
| Neuropathy | 934 | *22.59* | Neuropathy |
| Skin_disorders | 1055 | *25.51* | Skin disorder, Dry skin, Rash |
| Dyspepsia_Dysgeusia | 693 | *16.75* | Dyspepsia, Dysgeusia |
| Myalgia_Arthralgia | 754 | *18.24* | Myalgia, Arthralgia |
| Infection | 762 | *18.43* | Infection, Urinary tract infection, Vaginal candidiasis, Candidiasis, Cystitis, Catheter-related infection |
| Nail_disorders | 335 | *8.10* | Nail disorder, Onychoclasis, Onychalgia, Nail discolouration |
| Concomitants_Temperature | 280 | *6.78* | Hot flush, Flushing |
| Oedema | 278 | *6.73* | Oedema peripheral, Lymphoedema, Fluid retention, Oedema |
| Concomitants_Eye | 530 | *12.81* | Lacrimation increased, Foreign body sensation in eyes, Eye pain, Conjunctivitis, Dry eye, Eye irritation, Eye infection |
| Dyspnoe_Respiratory | 451 | *10.91* | Dyspnoea, Dyspnoea exertional, Lower respiratory tract infection, Cough, Upper respiratory tract infection |
| Pharynx_Larynx_Nares | 270 | *6.52* | Nasopharyngitis, Pharyngolaryngeal pain, Pharyngitis, Rhinorrhoea |
| Vein_disorders | 671 | *16.24* | Thrombophlebitis superficial, Deep vein thrombosis |
| Pain_specified | 482 | *11.66* | Pain in extremity, Back pain, Pain, Joint swelling, Abdominal pain, Abdominal pain upper, Chest pain, Headache, Ear pain |
| Mucosal_inflammation | 1873 | *45.30* | Mucosal inflammation |
| othertox | 1482 | *35.84* | includes all other listed toxicities (excluding the Haematological AES), which were experienced by less than 0.5% of the study population |

*Supplementary Table 2: Toxicity groups in TACT2*

| TACT2 - toxicity grouping | N= | % | |
|---|---|---|---|
| All patients | 1238 | *100* | |
| Alopecia | 1151 | *92.99* | Alopecia |
| Lethargy_Depression_Anxiety | 1094 | *88.35* | Fatigue, Insomnia, Depressed mood, Depression, Anxiety, Dizziness, Palpitations |
| Nausea_Vomiting | 852 | *68.79* | Nausea, Vomiting |
| Stomatitis | 830 | *67.03* | Stomatitis, Dry mouth, Oral candidiasis, Oral herpes |
| Constipation | 586 | *47.35* | Constipation, Haemorrhoids |
| Diarrhoea | 572 | *46.18* | Diarrhoea |
| Neuropathy | 198 | *16.03* | Neuropathy peripheral, Peripheral sensory neuropathy |
| Skin_disorders | 405 | *32.73* | Skin disorder, Dry skin, Skin infection, Localised infection, Wound infection, Rash |
| Dyspepsia_Dysgeusia | 317 | *25.62* | Dyspepsia, Dysgeusia, Abdominal distension, Duodenogastric reflux, Anorexia, Decreased appetite |
| Myalgia_Arthralgia | 105 | *8.51* | Myalgia, Arthralgia |
| Infection | 144 | *11.61* | Vulvovaginal candidiasis, Urinary tract infection, Catheter-related infection, Cystitis, Candidiasis, Infection |
| Nail_disorders | 39 | *3.12* | Nail disorders |
| Concomitants_Temperature | 69 | *5.55* | Hot flush, Flushing, Influenza-like illness |
| Oedema | 49 | *3.93* | Lymphoedema, Oedema peripheral |
| Concomitants_Eye | 172 | *13.88* | Lacrimation increased, Eye pain, Foreign body sensation in eyes, Dry eye, Conjunctivitis, Eye pruritus, Eye irritation |
| Dyspnoe_Respiratory | 209 | *16.86* | Dyspnoea, Cough, Lower respiratory tract infection, Upper respiratory tract infection |
| Pharynx_Larynx_Nares | 154 | *12.43* | Oropharyngeal pain, Nasopharyngitis, Rhinitis, Epistaxis, Pharyngitis, Rhinorrhoea |
| Vein_disorders | 235 | *18.97* | Thrombophlebitis superficial, Thrombosis, Phlebitis, Vein disorder, Vein pain |
| Pain_specified | 192 | *15.54* | Pain in extremity, Headache, Back pain, Musculoskeletal pain, Migraine, Breast pain, Joint stiffness, Bone pain, Abdominal pain, Abdominal pain upper |
| Palmar_plantarsyndrom | 5 | *0.37* | Palmar-plantar erythrodysaesthesia syndrome, Erythema |
| othertox | 695 | *56.14* | includes all other listed toxicities (excluding the Haematological AES), which were experienced by less than 0.5% of the study population |

### *Supplementary Table 3: Correlation between toxicities in TACT2*

| **TACT2** | Alopecia | LDA | Nausea | Stomatitis | Constipation | Diarrhoea | Neuropathy | Skin | Dyspeps. | Myalgia | Infection | Nail | Temp. | Oedema | Eye | Dyspnoe | Pharynx | Veins | Pain | Palmar | other |
|---|---|---|---|---|---|---|---|---|---|---|---|---|---|---|---|---|---|---|---|---|---|
| **Alopecia** | 1 | | | | | | | | | | | | | | | | | | | | |
| **LDA** | **0.1636** | 1 | | | | | | | | | | | | | | | | | | | |
| **Nausea** | 0.0218 | **0.2594** | 1 | | | | | | | | | | | | | | | | | | |
| **Stomatitis** | **0.1322** | 0.233 | **0.1373** | 1 | | | | | | | | | | | | | | | | | |
| **Constipation** | **0.1023** | **0.2187** | **0.2174** | **0.2285** | 1 | | | | | | | | | | | | | | | | |
| **Diarrhoea** | 0.0272 | 0.2039 | **0.1391** | **0.1144** | -0.0158 | 1 | | | | | | | | | | | | | | | |
| **Neuropathy** | 0.0343 | **0.1361** | 0.0655 | **0.1274** | 0.0406 | **0.1331** | 1 | | | | | | | | | | | | | | |
| **Skin** | 0.081 | **0.1232** | 0.0507 | **0.1165** | 0.0781 | 0.0674 | **0.15** | 1 | | | | | | | | | | | | | |
| **Dyspeps.** | 0.0803 | **0.1725** | **0.1237** | **0.106** | **0.1391** | 0.015 | 0.0735 | 0.0968 | 1 | | | | | | | | | | | | |
| **Myalgia** | 0.0179 | 0.0891 | 0.0403 | 0.0644 | 0.0433 | 0.0494 | 0.0695 | 0.0275 | 0.0645 | 1 | | | | | | | | | | | |
| **Infection** | 0.0331 | 0.0918 | 0.0213 | 0.0781 | 0.0551 | 0.0816 | 0.0392 | 0.0613 | 0.0537 | 0.0038 | 1 | | | | | | | | | | |
| **Nail** | 0.0174 | 0.0328 | -0.016 | 0.0101 | -0.017 | 0.0178 | 0.0722 | 0.0526 | **0.1132** | 0.0227 | -0.0163 | 1 | | | | | | | | | |
| **Temperature** | 0.0426 | 0.0982 | 0.0599 | 0.0707 | 0.0147 | 0.0192 | 0.0843 | 0.0575 | 0.053 | 0.0932 | -0.047 | 0.0117 | 1 | | | | | | | | |
| **Oedema** | 0.0131 | 0.0385 | 0.0006 | 0.0407 | -0.0127 | 0.0201 | 0.054 | 0.0629 | 0.0184 | 0.0262 | -0.005 | 0.0388 | 0.0419 | 1 | | | | | | | |
| **Eye** | 0.0527 | **0.1226** | 0.0669 | 0.0855 | 0.06 | 0.026 | 0.0406 | 0.0667 | **0.1446** | 0.0359 | -0.0054 | 0.0117 | 0.0394 | 0.0133 | 1 | | | | | | |
| **Dyspnoe** | 0.0155 | **0.1557** | 0.0602 | 0.0804 | 0.0482 | 0.081 | 0.0516 | 0.016 | 0.0463 | 0.0887 | 0.0325 | 0.0683 | 0.0399 | 0.0313 | 0.0124 | 1 | | | | | |
| **Pharynx** | 0.0517 | **0.1334** | 0.0661 | **0.137** | **0.1005** | 0.0536 | 0.0905 | 0.0914 | **0.1454** | 0.0313 | 0.0584 | 0.0611 | 0.0595 | 0.02 | **0.1229** | **0.1147** | 1 | | | | |
| **Veins** | 0.0837 | **0.1559** | **0.1072** | **0.1568** | 0.0906 | **0.1287** | **0.171** | **0.1846** | **0.1521** | 0.0482 | 0.0387 | 0.0355 | 0.0712 | 0.0889 | 0.091 | 0.0699 | **0.1342** | 1 | | | |
| **Pain** | -0.0206 | **0.1146** | **0.153** | 0.0595 | 0.0664 | 0.0703 | 0.062 | 0.0181 | **0.1631** | **0.1509** | 0.0183 | 0.0037 | 0.0985 | 0.0647 | 0.0882 | 0.0205 | 0.077 | 0.0932 | 1 | | |
| **Palmar** | -0.0487 | 0.001 | -0.096 | -0.0728 | **-0.2077** | **0.1963** | **0.2171** | **0.1** | -0.0363 | -0.0218 | -0.0211 | **0.1117** | 0.01 | 0.0321 | -0.026 | -0.0508 | -0.0091 | 0.0192 | 0.0174 | 1 | |
| **other** | 0.0092 | **0.1159** | **0.1023** | 0.0989 | 0.0819 | 0.0921 | 0.0905 | 0.0646 | 0.0759 | 0.0603 | 0.0572 | 0.0122 | 0.0714 | 0.0764 | 0.0533 | 0.0727 | **0.1097** | 0.0801 | **0.1057** | -0.006 | 1 |

LDA- Lethargy_Depression_Anxiety; Nausea – Nausea_Vomiting; Skin – Skin disorder; Dyspeps. -Dyspepsia_Dysgeusia; Myalgia - Myalgia_Arthralgia; Nail - Nail_disorder; Temp. - Concomitants_Temperature; Eye - Concomitants_Eye; Dyspnoe - Dyspnoe_Respiratory; Pharynx - Pharynx_Larynx_Nares; Veins - Vein_disorders; Pain - Pain_specified; Palmar - Palmar_plantarsyndrom; other – other toxicities

### *Supplementary Table 4: Correlation between toxicities in TACT*

| **TACT** | Alopecia | LDA | Nausea | Stomatitis | Constipation | Diarrhoea | Neuropathy | Skin | Dyspeps. | Myalgia | Infection | Nail | Temp. | Oedema | Eye | Dyspnoea | Pharynx | Veins | Pain | Mucosal | other |
|---|---|---|---|---|---|---|---|---|---|---|---|---|---|---|---|---|---|---|---|---|---|
| **Alopecia** | 1 | | | | | | | | | | | | | | | | | | | | |
| **LDA** | 0.0079 | 1 | | | | | | | | | | | | | | | | | | | |
| **Nausea** | 0.0288 | **0.1484** | 1 | | | | | | | | | | | | | | | | | | |
| **Stomatitis** | **0.12** | **0.2236** | **0.1037** | 1 | | | | | | | | | | | | | | | | | |
| **Constipation** | 0.0389 | **0.1101** | **0.2167** | **0.1398** | 1 | | | | | | | | | | | | | | | | |
| **Diarrhoea** | 0.0102 | **0.1811** | 0.0955 | **0.1894** | 0.0331 | 1 | | | | | | | | | | | | | | | |
| **Neuropathy** | 0.0392 | **0.2253** | -0.0735 | **0.2273** | 0.0348 | **0.1525** | 1 | | | | | | | | | | | | | | |
| **Skin** | 0.0249 | **0.225** | -0.0123 | **0.2486** | 0.0341 | **0.1889** | **0.2595** | 1 | | | | | | | | | | | | | |
| **Dyspeps.** | 0.0428 | 0.0279 | 0.0864 | **0.1443** | 0.0762 | 0.0965 | 0.0171 | **0.1214** | 1 | | | | | | | | | | | | |
| **Myalgia** | 0.0238 | **0.1748** | **-0.1853** | **0.1521** | -0.0187 | **0.1091** | **0.3468** | **0.1701** | 0.0388 | 1 | | | | | | | | | | | |
| **Infection** | -0.0285 | **0.1287** | 0.0544 | **0.1313** | 0.0473 | 0.0748 | 0.0502 | 0.0995 | 0.0217 | 0.0333 | 1 | | | | | | | | | | |
| **Nail** | 0.0229 | **0.1503** | **-0.1144** | **0.1503** | -0.0058 | **0.1184** | **0.2911** | **0.2458** | **0.1018** | **0.2484** | 0.0182 | 1 | | | | | | | | | |
| **Temperature** | 0.0598 | 0.0855 | -0.0237 | -0.001 | 0.0095 | -0.0204 | 0.0496 | 0.0639 | 0.0448 | 0.0158 | -0.0076 | 0.0232 | 1 | | | | | | | | |
| **Oedema** | -0.0097 | 0.0815 | -0.0416 | 0.0946 | 0.0302 | **0.1059** | **0.1834** | **0.1303** | 0.0924 | **0.1257** | 0.0048 | **0.1555** | 0.0013 | 1 | | | | | | | |
| **Eye** | 0.044 | 0.0718 | 0.0308 | 0.0923 | 0.0709 | **0.121** | 0.077 | 0.0987 | **0.1387** | -0.0044 | -0.008 | 0.0956 | 0.0258 | 0.0554 | 1 | | | | | | |
| **Dyspnoe** | 0.0234 | 0.0888 | 0.0216 | 0.0364 | 0.0034 | 0.0717 | 0.0568 | 0.0799 | 0.0694 | 0.0464 | 0.0374 | 0.0907 | 0.0886 | 0.0318 | 0.0704 | 1 | | | | | |
| **Pharynx** | 0.0114 | -0.0048 | -0.0005 | 0.0715 | 0.0167 | 0.0276 | 0.0493 | 0.0601 | 0.0698 | -0.0124 | **0.1455** | 0.0062 | 0.003 | 0.0588 | 0.0361 | 0.0925 | 1 | | | | |
| **Veins** | -0.0363 | **0.172** | **0.1029** | 0.0694 | 0.058 | 0.0481 | 0.0213 | 0.0486 | 0.0366 | 0.0266 | 0.0328 | 0.0074 | 0.0352 | 0.001 | 0.0094 | 0.0518 | 0.0012 | 1 | | | |
| **Pain** | 0.04 | **0.1158** | 0.0089 | 0.076 | 0.0431 | 0.0986 | **0.1415** | **0.1264** | 0.0861 | **0.1196** | 0.0351 | **0.1031** | 0.042 | **0.1071** | **0.1014** | **0.1129** | 0.0576 | 0.048 | 1 | | |
| **Mucosal** | 0.0408 | **0.2121** | 0.0576 | **0.487** | 0.0911 | **0.1638** | **0.2402** | **0.2423** | **0.1329** | **0.1722** | **0.1192** | **0.1945** | -0.0161 | **0.1138** | 0.093 | 0.0651 | 0.0456 | 0.0642 | 0.0763 | 1 | |
| **other** | 0.0504 | **0.1761** | 0.0437 | 0.0845 | 0.0135 | 0.0647 | 0.1149 | **0.1866** | **0.1086** | 0.0976 | 0.0324 | **0.1121** | **0.1205** | 0.0698 | 0.0734 | 0.0973 | 0.0508 | 0.069 | **0.1195** | 0.0779 | 1 |

LDA- Lethargy_Depression_Anxiety; Nausea – Nausea_Vomiting; Skin – Skin disorder; Dyspeps. -Dyspepsia_Dysgeusia; Myalgia - Myalgia_Arthralgia; Nail - Nail_disorder; Temp. - Concomitants_Temperature; Eye - Concomitants_Eye; Dyspnoe - Dyspnoe_Respiratory; Pharynx - Pharynx_Larynx_Nares; Veins - Vein_disorders; Pain - Pain_specified; Mucosal- Mucosal inflammation; other – other toxicities
